# Clinical validation of a point-of-care antibody test for COVID-19

**DOI:** 10.1101/2020.12.16.20248303

**Authors:** Monila Patel, Yogesh Lakhotia, Sneha Shah, Nilay Suthar, Cherry Shah, Sankarrao Kola, Radha Rangarajan

**Affiliations:** Department of Medicine, Smt NHL Municipal Medical College, Ahmedabad, India; Centre for Cellular Molecular Biology, Hyderabad, India; HealthCubed India Private Limited, Bangalore, India

**Keywords:** Covid-19, Antibody Test, Seroprevalence, Product Evaluation, Clinical study

## Abstract

The objective of this study was to evaluate the performance of a lateral flow antibody test for COVID-19, approved for use in India. Although many point-of-care antibody tests are available globally, they have been subjected to limited clinical validation. This has led to suboptimal outcomes in the field, where antibody tests play a significant role in tracking the immunity of individuals and communities. In this study an antibody test, ImmunoQuick that recognizes antibodies to the Nucleocapsid and Spike proteins of SARS CoV-2 was tested in 100 symptomatic patients with a positive or negative diagnosis of COVID-19, based on RT-PCR results. The overall sensitivity of the test was found to be 86.1% (95% CI: 76.4% to 92.8%) and specificity 100% (95% confidence interval: 73.5% to 100%). The sensitivity reached a peak of 95.7% with samples taken 17 days after the onset of symptoms. Overall, the sensitivity and specificity of the test are sufficient for assessing seroprevalence.

## Introduction

The COVID-19 pandemic is expected to persist beyond 2020, despite global efforts to contain the infection. Tracking the extent of spread is one of the key ways in which policy decisions regarding the opening of businesses and educational institutions can be made. This involves measuring the exposure of individuals to the virus, assessing protection from reinfection and monitoring immunity at the population level. Point-of-care (POC) antibody tests are well suited for this purpose; their cost and ease of use allow for repeated testing in clinical and non-clinical settings. Several POC antibody tests based on lateral flow immunochromatography have been approved for use globally (1). However, not all tests are backed by robust validation data. Hence, there have been questions about their sensitivity and specificity when deployed in the field (2, 3, 4, 5). In India, one of the earliest tests approved was from ImmunoScience India Private Limited. We undertook a clinical study to confirm the performance of ImmunoScience’s lateral flow immunoassay, ImmunoQuick, that detects IgG and IgM antibodies against the Nucleocapsid and Spike proteins of the SARS CoV-2 virus, from capillary blood (6). The study was conducted in a cohort of individuals with influenza-like symptoms, who were hospitalized at the Sardar Vallabhbhai Patel Institute of Medical Science and Research, Ahmedabad in the state of Gujarat. The objective of the study was to measure the sensitivity and specificity of the assay in individuals who had a confirmed diagnosis of being positive or negative for COVID-19. The data showed an overall sensitivity based on the detection of IgG and IgM antibodies as 86.1% (95% CI: 76.4% to 92.8%) and specificity 100% (95% confidence interval: 73.5% to 100%). These performance characteristics are sufficient for tracking and monitoring the immunity in individuals.

## Materials and Methods

The study was conducted between 4 Jul 2020 and 18 Jul 2020 and was registered with the Clinical Trials Registry of India with the registration number, CTRI/2020/07/026469. The study was approved by the ethics committee of Smt NHL Municipal Medical College, where the M.P serves on the Faculty. The college is affiliated to the Sardar Vallabhbhai Patel Institute of Medical Sciences and Research (SVPIMSR) where patients are admitted and treated. During the duration of the study, the hospital was a designated center for treating COVID-19 patients. The protocol was to admit anyone with influenza-like symptoms and conduct the Reverse Transcriptase-Polymerase Chain Reaction (RT-PCR) test. Those with a negative result were discharged while those with a positive result were kept hospitalized until 10 days past the onset of symptoms or until the resolution of symptoms.

A total of 100 subjects were enrolled in the study, 80 positive and 20 negative for SARS CoV-2 by RT-PCR. All subjects were above the age of 18. Those who had tested positive were at least 10 days past the onset of symptoms at the time of enrollment. Those who were negative by RT-PCR were mostly between day 3 to 9 from the onset of symptoms (14/20); 4 were between 1 to 2 days and 2 were between 16 to 17 days from the onset of symptoms. Individuals who were critically ill or had a history of immunosuppression or were participating in any other clinical trial were excluded.

### Study design

Informed consent was taken from eligible individuals in writing. Basic demographic information, medical history and RT-PCR results were recorded. Then the lateral flow antibody test was performed by lancing the index finger and applying two drops of blood in the sample window of the test cassette, followed by two drops of buffer, as per the manufacturer’s instructions. Results were read between 15 and 20 minutes after the application of blood. If individuals had a negative RT-PCR result but were positive by the lateral flow assay, venous samples were taken and frozen. These samples were then processed using an Enzyme-linked immunosorbent assay (ELISA) kit for total antibodies (COVIDscreen Plus) from Trivitron Healthcare Pvt. Ltd. A schematic view of participant flow is shown in Figure 1.

**Figure 1:**
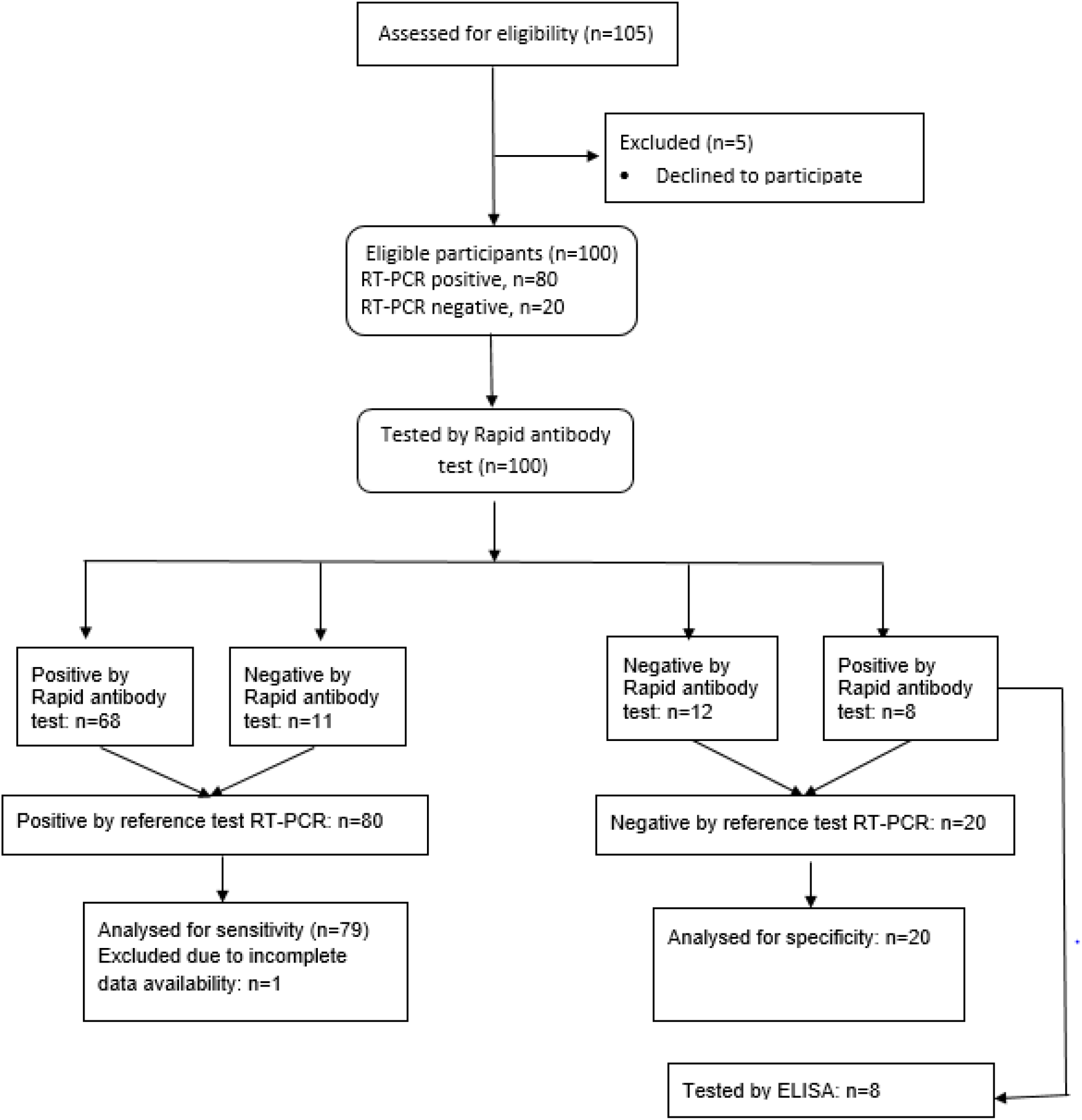
Participant flow diagram

### Data Analysis

A comparison of the results of the lateral flow assay and RT-PCR results was undertaken. The calculation of sensitivity and specificity were done as per the following formulae:

Sensitivity: TP/(TP+FN) where TP = True Positive; FN = False Negative

Specificity: TN/(TN+FP) where TN = True Negative; FP = False Positive

## Results

Individuals enrolled in the study were spread out between 10 to 73 days from onset of symptoms (Figure 2). The median was 15 days. Among patients who tested positive by RT-PCR, fever, breathlessness, and cough were the top 3 reported symptoms with an occurrence of 86%, 48% and 47% respectively. Similarly, hypertension and diabetes were the most reported medical history by the patients with an occurrence of 46% and 30% respectively.

**Figure 2:**
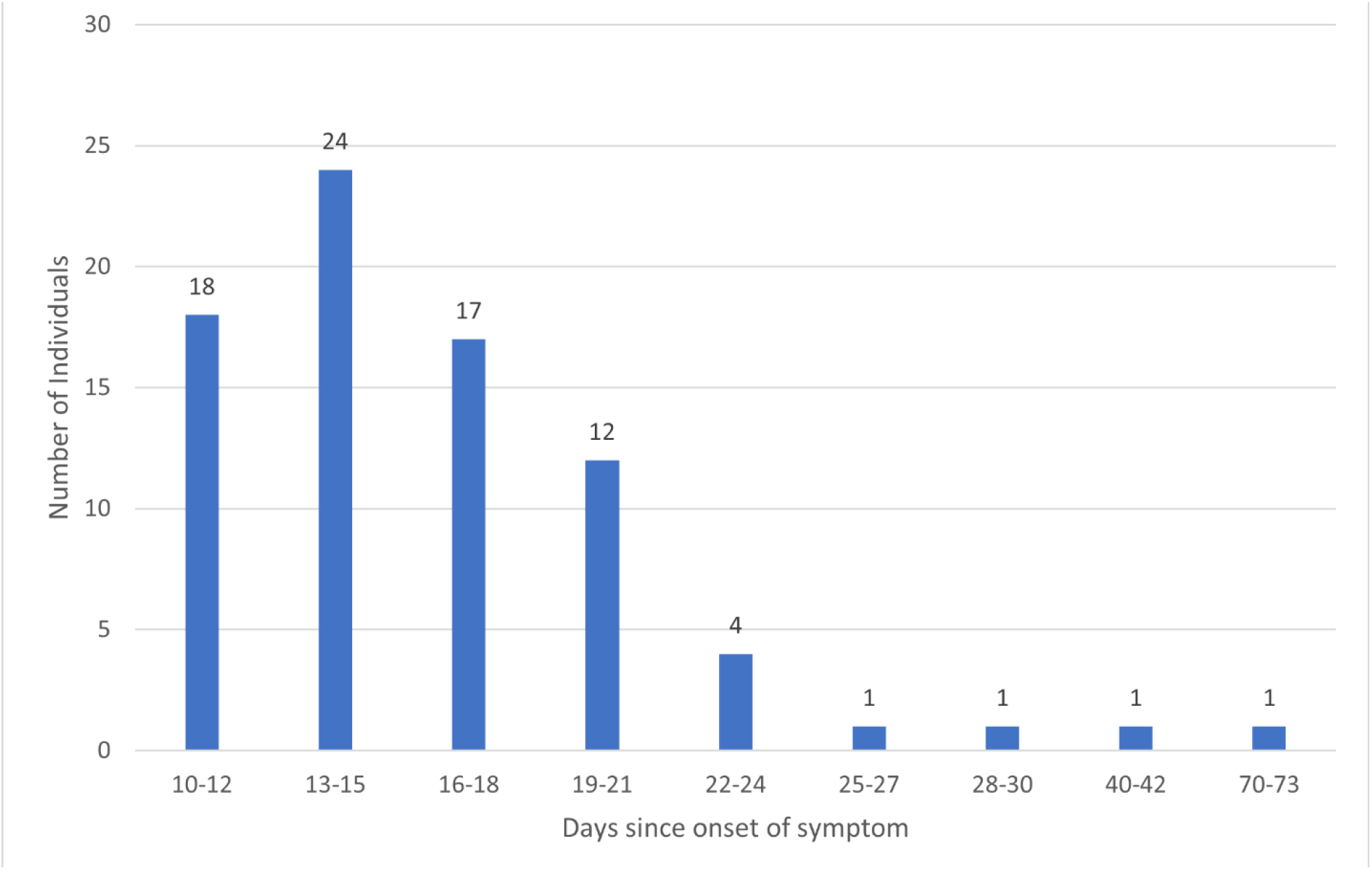
Distribution of samples by days from onset of symptoms

### Assay sensitivity

Of the 80 RT-PCR positive individuals, complete data were available for 79 individuals. 68/79 individuals were positive for either IgG or IgM antibodies by the Immunoscience lateral flow assay (Table 1). The sensitivity was calculated to be 86.1%, with a 95% confidence interval (CI) of (76.4% to 92.8%).

**Table 1:**
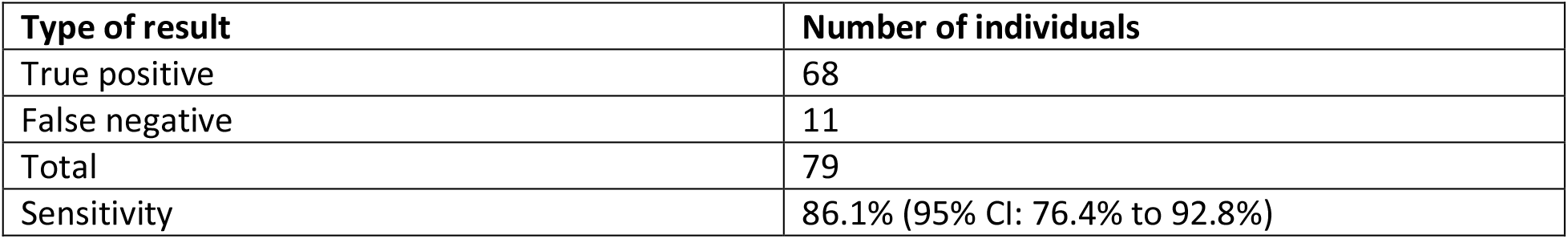
Sensitivity of assay

An analysis of sensitivity based on the number of days from onset of symptoms showed time dependent changes. When considering IgM positivity alone, sensitivity improved from 65.5% to 84.6% and dropped to 69.6% for samples taken at 10-13 days, 14-17 days, and 18-40 days respectively from the onset of symptoms. This trend is consistent with reported data on the rise and fall of IgM levels in patients, following a COVID-19 infection (7). When considering IgG positivity alone, sensitivity improved from 50.6% to 80.8% to 91.3% for samples taken at 10-13 days, 14-17 days, and 18-40 days respectively from the onset of symptoms. Finally, when considering overall sensitivity (IgM or IgG positivity), sensitivity improved from 75.9% to 82.3% to 95.7% for samples taken from patients at 10-13 days, 14-17 days, and 18-40 days respectively from the onset of symptoms (Figure 3). (A single data point from a patient at day 73 and negative for IgM and IgG antibodies was excluded). These data suggest that the sensitivity of the assay depends on the number of days from onset of symptoms. Highest sensitivity is achieved 17 days after the onset of symptoms.

**Figure 3:**
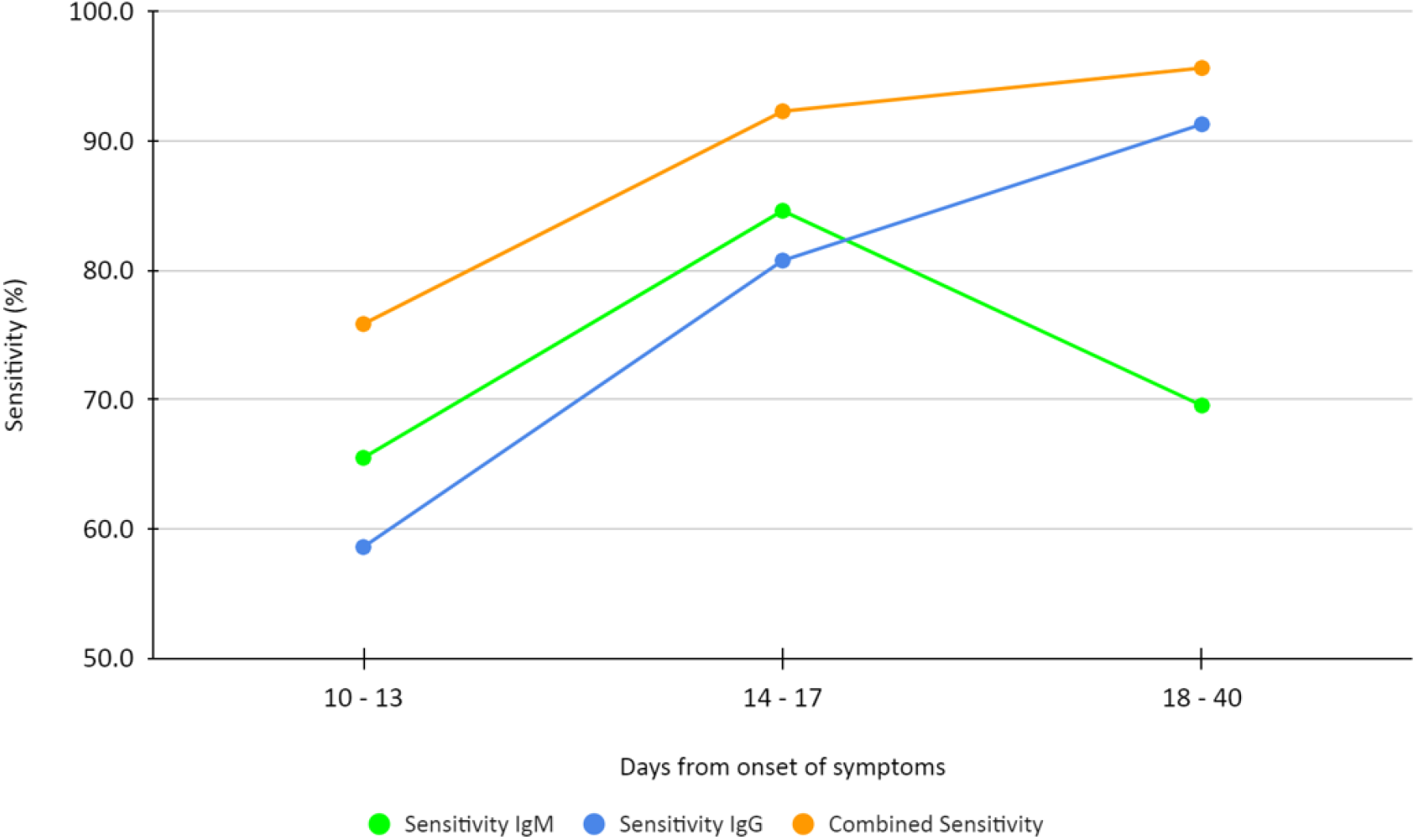
Test sensitivity with respect to days from onset of symptoms

In this study, we noted 11 false negative results. 9 out of 11 samples were taken between 10 and 15 days from the onset of symptoms, one at 19 and one at 73 days. It is possible that the antibody titer in these individuals was below the limit of detection of the lateral flow assay.

### Timeline of IgM and IgG responses

In this study, individuals were tested only once. Thus, a time course of antibody responses could not be derived. However, we analyzed the relationship between the number of days from the onset of symptoms and the type of antibody positivity (Table 3). The number of RT-PCR positive samples that were positive for IgM alone was 9, positive for IgG alone were 11 and positive for IgM and IgG were 48. The median time to IgM (alone) positivity was 12 days, IgM and IgG positivity was 16 days and to IgG positivity was 18 days, with overlapping ranges for the three types of immune profiles. Most individuals were positive for IgM and IgG antibodies, which is consistent with the time course of antibody responses reported in the literature (7).

**Table 2:** Changes in assay sensitivity vis-à-vis days from onset of symptoms

| Days from onset of symptoms | Number of samples | Number positive for IgM | Sensitivity (IgM alone) | Number positive for IgG | Sensitivity (IgG alone) | Number positive IgM or IgG | Overall sensitivity (IgM or IgG) |
| --- | --- | --- | --- | --- | --- | --- | --- |
| 10-13 | 29 | 19 | 65.5% | 17 | 58.6% | 22 | 75.9% |
| 14-17 | 26 | 22 | 84.6% | 21 | 80.8% | 24 | 82.3% |
| 18-40 | 23 | 16 | 69.6% | 21 | 91.3% | 22 | 95.7% |

**Table 3:** IgM, IgG positivity vis-à-vis days from onset of symptoms

|  | **IgM alone positive** | **IgG alone positive** | **IgM and IgG positive** |
| --- | --- | --- | --- |
| Number of samples | 9 | 11 | 48 |
| Median days since onset of symptoms | 12 | 18 | 16 |
| Range | 11 - 40 | 11 - 23 | 10 - 30 |

### Assay specificity

Of the 20 samples that were negative by RT-PCR, only 12 were negative by the lateral flow assay as well (Table 4). 8 samples, taken between days 1 to 9 from onset of symptoms, showed distinct IgM positive signals (Figure 4). As per the study protocol, venous blood from these individuals was analyzed by ELISA. All 8 were positive for antibodies (data not shown), taking the specificity to 100%, with a 95% confidence interval of 73.5% to 100%.

**Table 4:** Specificity of the assay

| **Type of result** | **Number of samples** |
| --- | --- |
| True negative | 12 |
| Positive by Lateral flow assay and ELISA | 8 |
| Total | 20 |
| Specificity | 100% (95% CI: 73.5% to 100%) |

**Figure 4.**
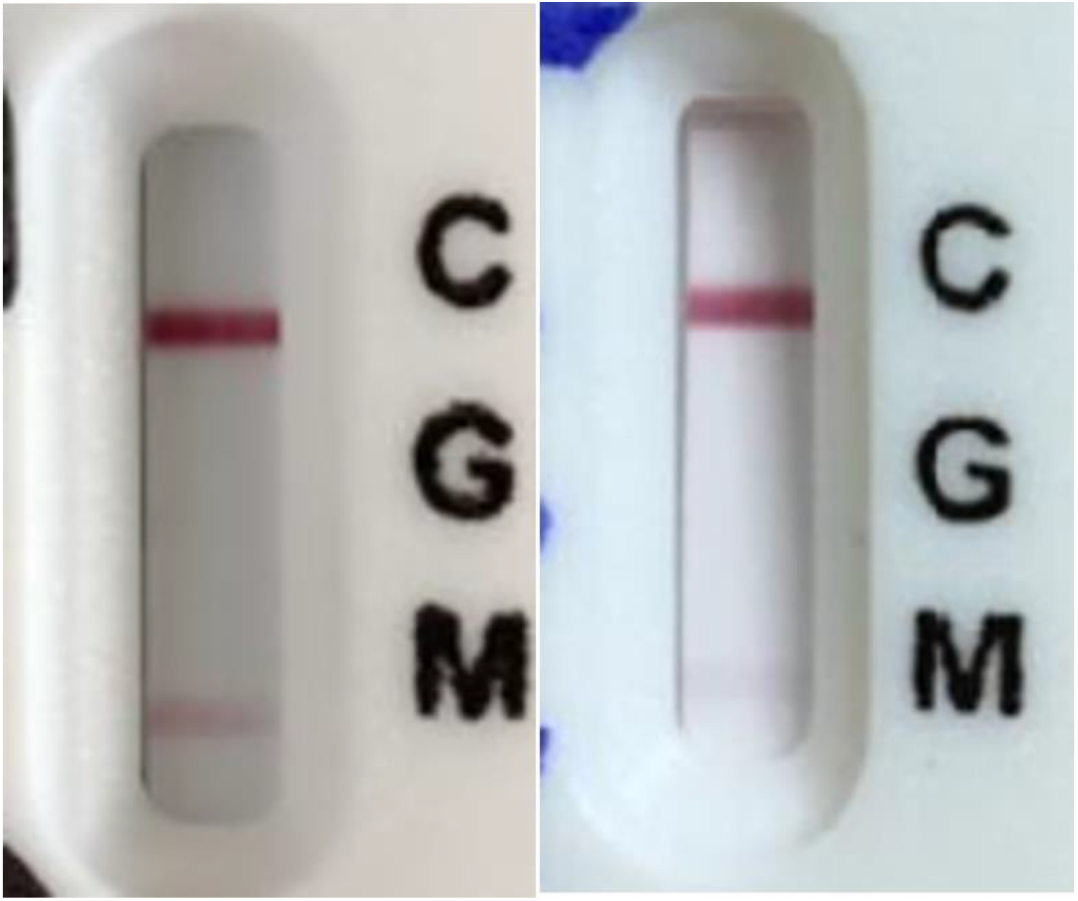
Images of 2 cassettes that were positive by the lateral flow assay and negative by RT-PCR. (C = Control, G = IgG, M= IgM)

## Discussion

The COVID-19 pandemic has brought into focus the need for simple test methodologies to track the immunity of individuals and communities. Antibody tests play an important role in this regard and are particularly valuable when available as POC tests.

In this study we evaluated ImmunoScience’s lateral flow immunochromatography assay for the detection of IgM and IgG antibodies specific to SARS CoV-2. We found the specificity to be 100% (95% CI: 73.5% to 100%), while the sensitivity was 86.1% (95% CI: 76.4% to 92.8%). Peak sensitivity of 95.7% was achieved after day 17. These results suggest that the test adequately serves the purpose of identifying those who have been exposed to the virus.

Amongst the RT-PCR positive samples correctly identified by the lateral flow assay, we found that the majority of the samples were positive for IgM and IgG consistent with the timing of study and the time course of antibody responses reported by others (7). We noted that there were 11 RT-PCR positive samples that were negative by the lateral flow assay. As the study did not involve any independent assessment of these samples by ELISA, it is assumed that these are false negative results. However, several recent reports have raised the possibility of T-cell mediated immunity (8). In future studies, such discrepancies must be addressed using alternate methods.

An analysis of results for the RT-PCR negative individuals revealed that 8 had antibodies to SARS CoV-2. While the reasons for the negative RT-PCR results cannot be pinpointed, we speculate that the timing of the test or suboptimal sample collection may have resulted in a false negative (9, 10). These results suggest that employing a multiplicity of tests may be appropriate when diagnosing symptomatic individuals.

The size of this study was small, n=99, with a 4:1 ratio of RT-PCR positive and negative individuals. The small size allowed the study to be completed in a short period of time and the higher number of RT-PCR positive individuals made it possible to probe the sensitivity of the assay in fuller detail. However, only a limited evaluation of specificity was possible with this study design. At the time of initiating the study, this was an acceptable trade-off as the manufacturer had claimed a specificity of 96% (oral communication) and the need to verify sensitivity was more urgent. Unexpectedly, the number of true negative individuals dropped from 20 to 12, greatly limiting the assessment of specificity. Future studies should include a larger number of true negative samples, characterized by RT-PCR and an ELISA or Chemiluminescence Immunoassay (CLIA) prior to enrollment in the study.

Overall, the data from this study support the use of the ImmunoQuick rapid test for serosurveillance.

## Data Availability

The data referred to in the manuscript was collected in the clinical study. No external data is being published in the manuscript.

## Acknowledgements

We thank Dr. N. Madhusudhana Rao from the Center for Cellular and Molecular Biology for helping us with the ELISA tests on the venous samples.

